# Knowledge, attitudes and usage of electronic medical records system at Ocean Road Cancer Institute in Tanzania; An example of EMR user satisfaction in sub-Saharan Africa

**DOI:** 10.1101/2023.09.18.23295750

**Authors:** Khadija Msami, Ugochukwu Okoroafor, Mohammed Mbwana, John Ngowi, Charles Lwanga, Habibu Luziga

## Abstract

**Background:** Electronic medical records (EMR) is a useful tool to facilitate workflow and improve the quality of patient care and patient safety. Ocean Road Cancer Institute (ORCI) started the computerization of patients’ records and running of administrative tasks via a customized system.

**Broad objective:** This study was designed to assess the extent of utilization, staff knowledge, and attitudes towards the EMR system at ORCI. A quantitative data collection tool was adopted and distributed.

**Methodology:** The study was conducted at Ocean Road Cancer Institute (ORCI), a national tertiary centre located in Dar es Salaam, Tanzania. Of the 259 employees, a standard structured tool for data collection was applied to the 154 staff calculated sample size.

**Results:** All the respondents (52) had a working experience of more than three months and had access to the EMR with 49 (94.2%) involved in direct patient care. 25(48.1%) were male and 27(51.9%) were female with a computer literacy rate of 92.3% (48). Only 2(3.8%) of the respondents held postgraduate degrees, and 25 (48.1%) each had diploma (college) and degree (university) level of education. 26(50%) felt the EMR was efficient and met user demands, 19(36.5%) felt it efficient but needing additional modification and only 4 (7.7%) felt it a wastage of time. Attitudes for the usage were fairly distributed with most claiming either slight increase (21, 40.4%) or increase (19,36.5%) quality of work but only 51.9% (27) felt the tasks became easier. 33(63.5%) of the respondents always use paper-based records together with EMR. Whereas 25(48.1%) felt the system provided precise information and 37(71.2%) agreed that the contents provided in the system met their needs, 15(28.8%) felt that the contents never met their needs and 27(51.9%) of the respondents felt that it is not precise. While majority (85%) overall were satisfied with the system and 31(59.6%) rating it as good. 5(9.6%) rated poor satisfaction and there was an even divide on finding the system complicated.

**Conclusions:** Overall, hospital staff know the potential benefits of proper implementation and use of an EMR in capturing clinical care. However, without proper and continuous ICT support, the EMR will continue to struggle with its adaptability and user satisfaction.

## Introduction

Electronic health (eHealth) is the utilization of information and communication technologies (ICT) to improve health systems and health service delivery by enhancing electronic flow of information (WHO, 2012). eHealth tools have been widely adopted in developed nations where its integration has led to improvement in their health care delivery.

EMR systems are electronic record of health-related information on an individual that can be created, gathered, managed, and consulted by authorized clinicians and staff within one health care organization. Provided for on a multitude of platforms run by various vendors, EMR systems can collect, store and make available to all members of the patient care team all relevant patient-care related information. EMR provides a clinical practice superior to paper-based records due to the ease of access, completeness, and legibility of documentation. Furthermore, EMR facilitates handovers of patients to staff during shift changes, improves the monitoring of revenue, reduces medical errors, facilitates easier prescription and dosing, and significantly reduces costs of healthcare delivery (James Menke, 2001). Although used synonymously with electronic health records (EHR), EHR can be considered an extension of EMR where there is exchange of medical information between health facilities and other stakeholders (Garets& Davis, 2006).

EMR systems have been around for a long time (Atherton, 2011), however low-income countries continue to experience difficulties with adoption and management of the systems due to resources and absence of foundation for implementation. The WHO explains that for its adoption, eHealth requires a purposeful action by nations to mobilize existing resources and plans that foster a base for sustainability in improving efficiency in the health sector. Others have found success with scaling up the program from experiences of pilot projects.

Ocean Road Cancer Institute (ORCI), adopted an EMR system in 2012, nicknamed ‘*Niara*’. In 2015 after undergoing some modifications, customization, and addition of features to suit the institute’s needs, the EMR system developed a more comprehensive outlook and was renamed ‘*Inaya*’. *Inaya* included different modules encompassing collection of patient data during clinical consultations, diagnoses and pharmacy prescription dispensation. The system modules also include administrative activities such as billing and accounting, budget, procurement, storage and human resources management services.

As a component of eHealth systems, electronic health records (EHR) play a fundamental role in patient management and effective medical care services. Thus, the adoption of EHRs in regions with a lack of infrastructure, untrained staff, and ill-equipped health care providers is of paramount importance. However, main barriers to adopting EHR software in low- and middle-income countries include the cost of its purchase and maintenance, incompetent and under-developed systems, lack of technical expertise as well as knowledge attitudes and practices of the users of these systems (WHO,2012). Despite this, data shows that the cost of healthcare is inevitably reduced with implementation of an EHR (Blumenthal, 2009; Bar-Dayan et al., 2013).

As ORCI becomes accustomed to this relatively new technology, there is a need to determine not only its usefulness in improvement of care and service delivery, but also staff perception of its implementation. This will lead to recommendations for features and components currently omitted from the *Inaya* system. This study was the first of its kind at ORCI and aims at filling the knowledge gap of user perceptions towards EMR and to assess its interoperability across departments involved in patient care. The ensuing results will describe the extent of functions and scope of usage of the system, key information for future applications and upgrades to the system. Indeed, making a system perfect in developing countries could still be an insurmountable task, but aspiring to have a flawless system through evidence-based recommendations is a positive step that will increase knowledge, skills, efficiency, and positive perceptions of EMR.

Decision and policy makers understand that technology stands to improve and have positive utility to healthcare workers in the context of patient care. This is exemplified by the inclination towards generation and usage of data and implementation of EMR systems nationwide. Through this research, the MoHCDGEC will receive a tool for evaluation of EMR that is customized for its hospitals implementing the EMR. This tool will aid the MoHCDGEC to conduct comparison evaluations across different (software) platforms currently being used at the different hospitals. This current study will highlight levels of satisfaction and perceived changes in quality of work among various consumer cadres at ORCI and whether this is associated with usage of system.

## Methodology

The study was conducted at Ocean Road Cancer Institute (ORCI), a national Tertiary healthcare centre located in Dar es Salaam, Tanzania. The target population included all eligible ORCI staff at the time of the study. Accordingly, ORCI has employed 259 employees by January 2018.

Using stratified probability sampling design with a standard structured tool for data collection, proportional allocations were used in assigning sample to desired categories. In the determination of the desirable sample size of the research, we assumed the level of precision to be ±5%, the confidence level to be 95% and the degree of variability will be set at 0.5 (P). The following formula was used for calculating the finite sample size from our population:

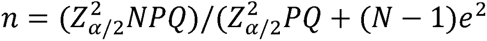

Whereas:

N – Is the population size
n – Is the sample size
Z – is the abscissa of the normal curve that cuts off an area α at the tailse
e – Desired level of precision
P - Is the estimated proportion of an attribute that is present in the population.

In this study we will assume that, e = 0.05, α = 0.05, P = 0.5, Z_α/2_= 1.96,

N= 259 = Number of all staff at ORCI by January 2018,

Calculations;

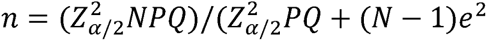

n= 248.74/1.61

*n= 154*

A sample of 154(59.5%) out of 259 eligible respondents was selected. Furthermore, investigators applied exclusion criteria to retain only employees that had direct patient interactions daily along with access to the EMR. As a result, only 78 eligible respondents were retained for this study. Of the 78, only 52 respondents returned the questionnaires on time.

The questionnaires assessed both user satisfaction and task-oriented frequency of use with some modifications to assess the effectiveness of EMR system at ORCI from the standpoint of the user. (Lærum & Faxvaag, 2004).

Upon standardization, the questionnaire was reviewed and translated by a physician external to the research team who was blinded to the initial version of the questionnaire. Furthermore, Likert scales were utilized to assess the knowledge, attitudes and level of satisfaction regarding EMR use among staff at ORCI.

To assess the validity of the questions in the questionnaires and whether the respondents were able to understand them without any hesitation, pre-testing of the questionnaires was done at ORCI, 20 randomly selected staff participated in the pre-test. The selected staff for pretest survey were excluded in main survey to avoid biasness of information.

### Conceptual framework of the study

The ethical clearance for the study was sought from the Research Ethics Committee at ORCI. Data analysis was carried out using SPSS version 23 and Microsoft excel 2013. Likert scale, bivariate analysis and multivariate regression analysis were employed to calculate rating averages, test categorical relationship between variables and estimate a regression model, respectively.

## RESULTS

The results of the analysis were summarized through descriptive statistics, rating analysis, bivariate analysis using the chi square method and the multivariate regression analysis.

A total of 52 respondents filled the questionnaire anonymously. The socio demographic profiles of the respondents are given in Table 3. The analysis determined that all respondents 52 (100%) had a working experience of more than three month. Most (28) staff were aged between 35-50 years with 53.8%. Among of the respondents, 25(48.1%) were male and 27(51.9%) were female. The interview was scattered throughout the cadres with 71.2% coming from Medical and Allied Health Science Service directorates. Among the respondents, 49(94.2%) were involved in direct patient care, while 3(5.8%) worked with patients indirectly as part of a care team. The level of education was distributed evenly for the staff with diploma (college) and staff with degree (university) at 25(48.1%) apiece and only 2(3.8%) of the respondents held postgraduate degree. The computer literacy rate amongst the respondent was 48(92.3%) and all of them 52(100%) have been locally trained and have access to the EMR at their working department.

**Table 1.**

| STAFF CADRE | ELIGIBLE | RESPONDENT |
| --- | --- | --- |
| Nurses | 27 | 25 |
| Radiotherapists | 14 | 7 |
| Medical Doctors | 13 | 6 |
| Pharmacists | 2 | 1 |
| Accounting officers | 3 | 3 |
| Procurement officers | 1 | 1 |
| Medical Records | 11 | 3 |
| Planning officers | 1 | 1 |
| Laboratory Technologists | 4 | 4 |
| Human resource officers | 2 | 1 |
| **TOTAL** | **78** | **52** |

**Table 2.**

| Independent variables | Dependent variables |
| --- | --- |
| Demographic characteristics of staff | Staff Attitudes regarding EMR |
| Profession of staff | Number of patients consulted per mode of record |
| Level of education | Completeness of data in EMR |
| Department | Accuracy of information entered in EMR |
| Availability of EMR at department |  |
| Computer literacy |  |
| Knowledge regarding EMR |  |

**Table 3.**

| Variable | Category | Respondents |
| --- | --- | --- |
|  |  | (n=52) |
| Gender | Male | 48.1 |
|  | Female | 51.9 |
|  | **Total %** | **100.0** |
| Age | < 35 | 44.3 |
|  | 35-50 | 53.8 |
|  | > 50 | 1.9 |
|  | **Total %** | **100.0** |
| Direct patient care | Yes | 94.2 |
|  | No | 5.8 |
|  | **Total %** | **100.0** |
| Working Experience<br>of more than 3 months | Yes | 100 |
|  | No | 0 |
|  | **Total %** | **100.0** |

### 4.0 Description of the respondents’ Attitude to the EMR

When assessing the attitude of the respondents for the efficient of the EMR installed at ORCI, 26(50%) felt it was efficient and meet user demands. 19(36.5%) claimed that the EMR was efficient but still need extra modification and only 4(7.7%) claimed that the EMR was wastage of time and need to be re-developed. Further analysis also revealed that 37(71%) of correspondents reported that the system was user friendly and easy to use.

### 4.1 Description of the respondent’s usage of EMR

On rating the success of the EMR installed, 44(84.6%) of the respondents had reported positively upon the usage of the EMR system installed at ORCI and this was either excellent, good, or fair as elaborated in the table 4 below.

**Table 4.** illustrating the success rate of the EMR system installed at ORCI departments.

|  |  | Frequency | Percent |
| --- | --- | --- | --- |
| Valid | No answer | 7 | 13.5 |
|  | Poor | 1 | 1.9 |
|  | Fair | 15 | 28.8 |
|  | good | 20 | 38.5 |
|  | excellent | 9 | 17.3 |
|  | Total | 52 | 100.0 |

**Table 5.**

|  |  | Frequency | Percent |
| --- | --- | --- | --- |
| Valid | No | 16 | 30.8 |
|  | yes | 36 | 69.2 |
|  | Total | 52 | 100.0 |

**Table 6.** shows satisfaction with *inaya*.

|  |  | Frequency | Percent | Valid Percent |
| --- | --- | --- | --- | --- |
| Valid | poor | 9 | 17.3 | 17.3 |
|  | satisfactory | 43 | 82.7 | 82.7 |
|  | Total | 52 | 100.0 | 100.0 |

Accordingly, to the usage of the system, 21(40.4%) respondents claimed that the quality of the department work has slightly increased, 19(36.5%) stated that the quality of their work increased. Due to the usage of the system 27(51.9%) respondents stated that the performance of their task has become easier.

### Analysis of ongoing usage of paper-based records together with EMR

33(63.5%) of the respondents attest to always use the paper based medical record or the chart summary as an information source on their daily clinical work. With only 7(13.5%) who stated that they never or almost never still use the paper based medical record or the chart summary as an information source on their daily clinical work. Likewise, 19(36.5%) always or almost always use EMR as an information source in their daily clinical work while 16(30.8%) about half of the occasions use EMR as an information source in their daily clinical work whereas 11(21.2%) never or almost never use EMR as an information source in their daily clinical work. While transferring patients related information to other person or instances (by printouts or by electronics transmission), 26(50%) respondents agreed on their use of EMR for this purpose, 8(15.4%) use EMR half of the occasion for transferring patients related information and about 13(25%) never use EMR for the purpose.

On the preciseness of the information provided by the system, 27(51.9%) of the respondents felt that it does not provide as much precise information as they would hope a system would, whereas a combined tally of 25(48.1%) felt the system provided precise information. 15(28.8%) felt that the contents never met their need compared to a tally of 37(71.2%) who agreed that the contents provided in the system met their need, hence they were satisfied content wise.

### Assessment of EMR accuracy at ORCI

Following our analysis, we were able to obtain information that suggests that 36(69.2%) of all respondents reported that the EMR system was accurate while 16(30.8%) suggested that the system was inaccurate and felt that developers must incorporates some new ideas for them to be fully satisfied with the system accuracy.

### Frequency table highlighting EMR accuracy at ORCI

Lastly, 23(44.2%) and 9(17.3%) agree and strongly agrees respectively that the EMR is worth the time and effort required to use it, where only 3(5.8%) disapprove that EMR is worth the time and effort required to use it. With majority in agreement with the system, 31(59.6%) rated EMR with good satisfaction which was the highest grading rate used in the questionnaire, while only 5(9.6%) gave a poor satisfaction rating to the EMR.

Overall, 85% of the users responded that they were satisfied with the system, because they were able to find all the necessary registers in the system, could track patients, do transfers and referrals, receive notifications on lab results, easy to learn, understand, verify data entry and reports, retrieve and share reports, good for case management, web based, useful in day-to-day work and training on the system was satisfactory. The outstanding 15% were left unsatisfied with the EMR because they needed more clinical practice to be conversant with the system, system did not have all the validations necessary for quality data entry, difficult to navigate and involved many clicks.

In relation to satisfaction, although many users agreed that the system was to their satisfaction, when asked whether it was complicated, they were equally divided in response; 51% felt it was complicated to use and the rest felt it was not.

## DISCUSSION

Traditionally the focus of research on technological advances in health care like the implementation of an EMR, has chiefly been on its design, practicability, and impetus for implementation (Anderson, 1997; Jha et al., 2006; Poon et al., 2006; Lorenzi et al., 2008). These studies have been disproportionately conducted by developed countries and have highlighted gaps in knowledge of how end users respond to the technology when newly adopted and in operation at the facility. This gap in health technology knowledge has been exacerbated by the design of the research typically used, chiefly behavioral models that explain human behavior through applications of theory and prediction (Chau& Hu, 2001; Schaik et al., 2002; Yi et al., 2006; Holden & Karsh, 2009). These behavioral studies that have used the technology acceptance model (TAM) whose constructs are adaptations of the theory of reasoned action, concluded that perception of ease of use of the system over that of usefulness of the system is given more weight by health care professionals (Holden & Karsh 2010; Al-Mujaini, 2011). That means to ensure that the system works, teasing out perceptions of healthcare staff on the system is of paramount importance.

EMR systems are notorious for failure, with global estimates showing up to 50% of EMR systems failing to be properly utilized (Willyard 2010). Once an EMR system is in place, its successful application lies in acceptance and utilization by health care staff (Berg, 1997). Healthcare providers in sub-Saharan Africa seeking to use EMR systems in their practice face many challenges, one of which is standardization of software of EMR systems. This remains a prerequisite for compatibility and interaction between health care facilities and this remains lacking particularly in sub-Saharan africa (Okoroafor, 2019). However, even where no interconnected eHealth systems are in place, a facility based EMR system that is easy to use and contains multi-modules to facilitate interdepartmental activities is a positive critical first step. After all, according to Ketikidis and colleagues (2012), implementation of more complex technologies with broad application is a difficult feat even for high income countries.

EMR implementation in Tanzania has been subject to some study. One (1) conducted at the Emergency Medicine Department in Muhimbili National (MNH) listed requirements for adequate implementation of EMR in low-income countries. *Inaya’s* implementation reflected the recommendations by the Muhimbili study and the system had further undergone several variations with documentation templates customized extensively to meet departmental needs. ORCI staff also were provided numerous trainings session and the institution employed three ICT personnel supplemented by a volunteer force. In contrast to the single day launch event (*Go Live*) described in the Muhimbili study, ORCI opted to have the existing paper medical records (PMR) in parallel to the gradual implementation of the EMR. This phased approach was due to the complexities of managing a tertiary oncology setting and balancing the hospital-wide approach over a single department. These unique requirements include oncology specific documentation, supportive tasks for care coordination, and automated systems and safety checks to complement a field with shortages in expertise. In high income countries, EMR systems vendors typically offer direct guidance to the end-user along with routine updates (3), a consideration not readily applied for low income countries (LIC).

A prequel study conducted at ORCI assessed the documentation rate in the EMR system, comparing it to that of the paper-based record. This study found superior rates of documentation of diagnostic and management information in the paper-based records highlighting a misapplication of the system? (Okoroafor et. al). This quantitative exploration revealed a need for an organizational solution. This led to some review of ORCI workflows and identify inefficiencies to guide customization of the EMR to fit departmental needs with 51 updates and 2 major integrations: increasing its capacity with automated hematology machines and including a patient SMS gateway.

Analyzing user satisfaction of EMRs was the logical next step after exploring the systems implementation, capacity, and use particularly during the adoption of EMR exclusively at ORCI. To the best of our knowledge, this study is the first of its kind investigating user satisfaction in real time use of EMR in Tanzania. This study has uncovered

Overall, the results suggest our study participants were very aware of the need for an EMR and the potential benefits to be gained from its fully functionality, with 84.6% of respondents reflecting positively on its usage. However, the system fell short of gaining trust as suggested by 51.9% of respondents saying the EMR does not always contain all the necessary information needed for clinical day-to-day practice. 63.5% of respondents saying they always use the paper records for their day-to-day practice.

The participant experiences also make it clear that the promised advantages were yet to be fully realized and may not be achievable without a different approach to user support, human resource, upfront investment and change management, spanning all deployment context. Similar potential benefits from EMR use, and barriers, have been observed with other EMR implementations in low resource settings [Ohabuwa 2012].

This study was not without its limitations. It was conducted in a small-time frame with a limited budget allowing for just a fraction of the eligible staff to participate in the study. Furthermore, being that this study was conducted at by the institution itself, there is a potential for biased responses by staff. This was overcome by supplementing the results with objective data collection methods like directly observing practice, ensuring confidentiality to study participant responses, and collection only de-identified data.

A major strength of this study is the use of a standardized questionnaire in assessing EMR utility. Its use quickly estimates the potential impact of the EMR system on health care delivery and helps identify problematic areas in clinical tasks for which the EMR system either is not used, or for which performing the task is more difficult when using the system. With continued standardized surveys like this one, we may be able to compare data across time, site, or vendors.

Overall, hospital staff know the potential benefits of proper implementation and use of an EMR in capturing clinical care. However, without proper and continuous ICT support, the EMR will continue to struggle with its adaptability and user satisfaction.

Future studies can explore factors that influence use and user satisfaction of EMRs based on the D&M and TAM models in order to determine the key factors that influence EMR implementation in sub-Saharan Africa.

## Data Availability

All data produced in the present work are contained in the manuscript

